# Integrative Analysis of GWAS and Single-Cell eQTL Data Identifies Immune Cell-Type-Specific Genetic Signals Co-occurring in Inflammatory Diseases and Lung Cancer Risk

**DOI:** 10.1101/2025.10.04.25337327

**Authors:** Langxuan Yu, Wen Zhe, Peng jin, Yanjie Niu, Liyan Jiang

## Abstract

**Background:** Non-small cell lung cancer (NSCLC) frequently co-occurs with chronic inflammatory conditions such as COPD, RA, IPF, psoriasis, and Dermatomyositis. While epidemiological links are established, the genetic basis of this co-occurrence in specific cell types remains poorly resolved.

**Methods:** We applied a signal-driven colocalization approach to NSCLC GWAS index variants (*p* < 1 × 10^−6^), integrating single-cell eQTL (sc-eQTL) data across cell types within ±100 kb windows to identify stable, compact variant sets that jointly explain GWAS and sc-eQTL signals. Loci with variant sets that optimally account for both NSCLC and sc-eQTL signals were evaluated for directional consistency between GWAS and sc-eQTL effects. In a subset of these loci, we performed locus-specific Mendelian randomization to evaluate whether the same set of genetic variants that influence risk of inflammatory comorbidities also show directionally concordant effects on NSCLC risk.

**Results:** Hundreds of genomic loci with robust concordance between NSCLC risk and cell type-specific gene regulation were identified. Directional analysis revealed consistent regulatory effects in specific immune contexts, and Mendelian randomization supported shared genetic liability between multiple inflammatory diseases and NSCLC risk.

**Conclusion:** Integrative analysis of GWAS and single-cell eQTL data enables the identification of shared regulatory architectures linking inflammatory diseases to lung cancer risk. This framework may support the role of immune cell-type-specific regulation in disease etiology and may inform strategies for risk stratification and mechanistic investigation.

## Background

Non-Small Cell Lung Cancer (NSCLC) is the leading cause of cancer death worldwide, accounting for approximately 18% of all cancer-related mortality[1]. Despite advances in early detection and therapy, over 60% of patients present with advanced or metastatic disease, and 5-year survival remains below 20%[2]. Prevention through risk factor modification may offer a more effective strategy to reduce global burden.

Recent efforts have turned to single-cell expression quantitative trait locus (sc-eQTL) data to pinpoint the cell types and target genes through which genetic risk variants operate[3]. However, current sc-eQTL resources are inherently fragmented: public releases usually typically report only variant–gene pairs with nominal p values below 0.05, while the majority of tested associations are omitted[3–9]. This selective reporting results in incomplete and sparse regulatory maps across cell types. Consequently, conventional colocalization methods such as COLOC, which assume dense SNP coverage, consistent linkage disequilibrium (LD) structure, and homogeneous variant representation across datasets, often yield inaccurate or irreproducible colocalization results when applied to NSCLC GWAS in conjunction with uncomplete sc-eQTL data[10,11].

Besides, Chronic inflammatory diseases including IPF, dermatomyositis, COPD, RA, and psoriasis are epidemiologically linked to increased lung cancer risk[12–16]. Chronic inflammation promotes carcinogenesis through immune activation and tissue remodeling. Mendelian randomization studies suggest potential causal roles, implicating shared genetic liability[17–21]. However, these analyses operate at the trait level and do not resolve specific variants or cell-type-specific regulatory mechanisms. the coarse granularity of Mendelian randomization analysis and sc-eQTL data motivate a shift from epidemiological observation to cell-type-resolved molecular mechanism. Although sc-eQTL data theoretically enable such resolution, their incompleteness risks propagating bias rather than clarifying causality.

To address these challenges, we introduce an integrative framework engineered to operate under fragmented genomic architectures. Leveraging high-resolution sc-eQTL data despite their sparsity, our approach prioritizes shared risk variants with cell-type-specific regulatory potential between NSCLC and chronic inflammatory diseases. By combining signal-driven variant selection[22], cross-modal concordance quantification, directional effect assessment[23,24], and locus-specific Mendelian

Randomization[25,26], our method identifies high-confidence loci where genetic effects on gene expression align with both NSCLC risk and inflammatory disease susceptibility, thereby bridging the gap between association and mechanism in the context of realistic, incomplete genomic data.

Anchoring on GWAS variants with suggestive association (p < 1 × 10^−6^), we integrate local sc-eQTL signals within ±100 kb windows and apply a signal-driven variant set prioritization procedure. This approach identifies compact and stable sets of variants that collectively explain the maximum shared genetic signal across GWAS and sc-eQTL modalities, explicitly accounting for the sparse and heterogeneous structure typical of public sc-eQTL resources. To ensure prioritized loci are relevant to NSCLC– inflammation comorbidity, we further filter them based on significant enrichment of associations in independent inflammatory disease GWAS summary statistics, serving as a phenotype-informed relevance screen.

## Method

### 1. Signal-Driven integration of GWAS and Fragmented sc-eQTL

Conventional colocalization methods (e.g., COLOC) assume complete SNP coverage, consistent linkage disequilibrium (LD) structure, and homogeneous variant representation across datasets, which is rarely satisfied in cross-study integration of single-cell eQTL (sc-eQTL) data[10,11]. To address this, we performed a signal-driven integrative variant prioritization framework designed to operate under the realistic constraints of fragmented single-cell eQTL data. For each candidate locus, defined as a ±100 kb genomic window centered on a GWAS lead SNP (p < 1 × 10^−6^),we include variants that satisfied all of the following criteria: (i) present in both GWAS and single-cell eQTL (sc-eQTL) summary statistics; (ii) reported as significant cis-sc-eQTLs (nominal p < 0.05 in the original sc-eQTL study); (iii) with LD estimates available from the 1000 Genomes Phase 3 European reference panel (1000G EUR). Overlapping loci were analyzed independently to prevent conflation of distinct regulatory architectures. All variants were harmonized to GRCh37 (hg19) using the UCSC liftOver tool (https://genome.ucsc.edu/cgi-bin/hgLiftOver). We queried the GRCh37 reference via the Ensemble REST API (https://grch37.rest.ensembl.org) and only retained variants with alleles consistent across datasets. Only variants with consistent allele coding and forward-strand alignment were retained for downstream analysis.

To quantify the degree of shared genetic signal between GWAS and sc-eQTL associations, we adopted a symmetric, energy-based heuristic termed the Signal Overlap Proportion (SOP). SOP is defined as the average of two directional ability metrics: the proportion of total GWAS association signal that explained by the selected sc-eQTL-derived causal variant set; and the proportion of total sc-eQTL signal explained by the GWAS-derived causal variant set.

Formally:

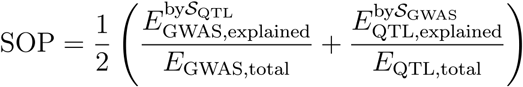

where each *E*_*explained*_ is computed as the cumulative signal energy accounted for by the selected variant set during the greedy conditional Z-selection procedure, and all energies are evaluated in the same spectrally truncated subspace to ensure comparability. We use SOP ≥ 0.7 as an empirical threshold to prioritize loci with substantial cross-modal signal overlap.

A complete description of the procedure is provided in the ***Supplementary Methods***. A theoretical justification of the proposed framework is provided in ***Supplementary Note 1***, including the statistical consistency of variance-normalized signal energy estimation, the validity of block-level LD definitions, the convergence of spectral regularization, and the stability under incremental SNP addition. By this design, our framework serves as an alternative in the presence of fragmented QTL data, thereby enabling inference in realistic genomic settings where traditional methods may lack reproducibility. Custom analysis scripts implementing the procedures described in this study are available at: https://github.com/SpaceRan/Signal-Driven-integration-of-GWAS-and-Fragmented-sc-eQTL. The analytical pipeline is illustrated in Figure 1

**Fig 1a.**
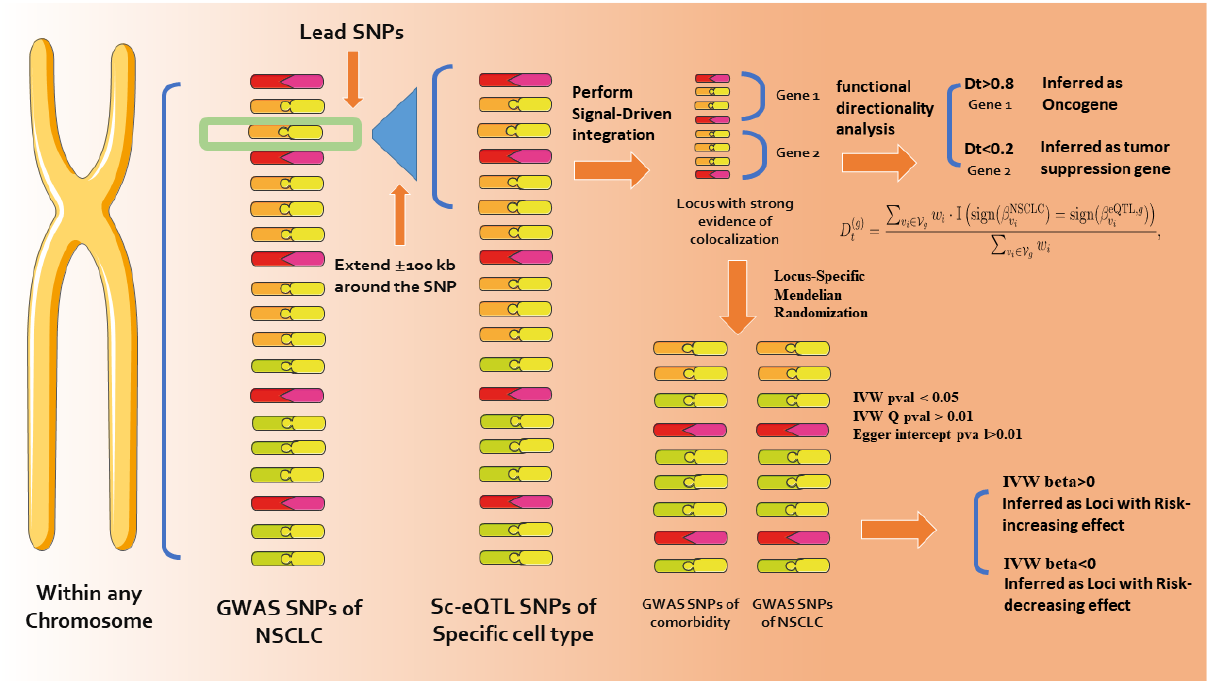
Workflow of the analytical pipeline.

**Fig 1b.**
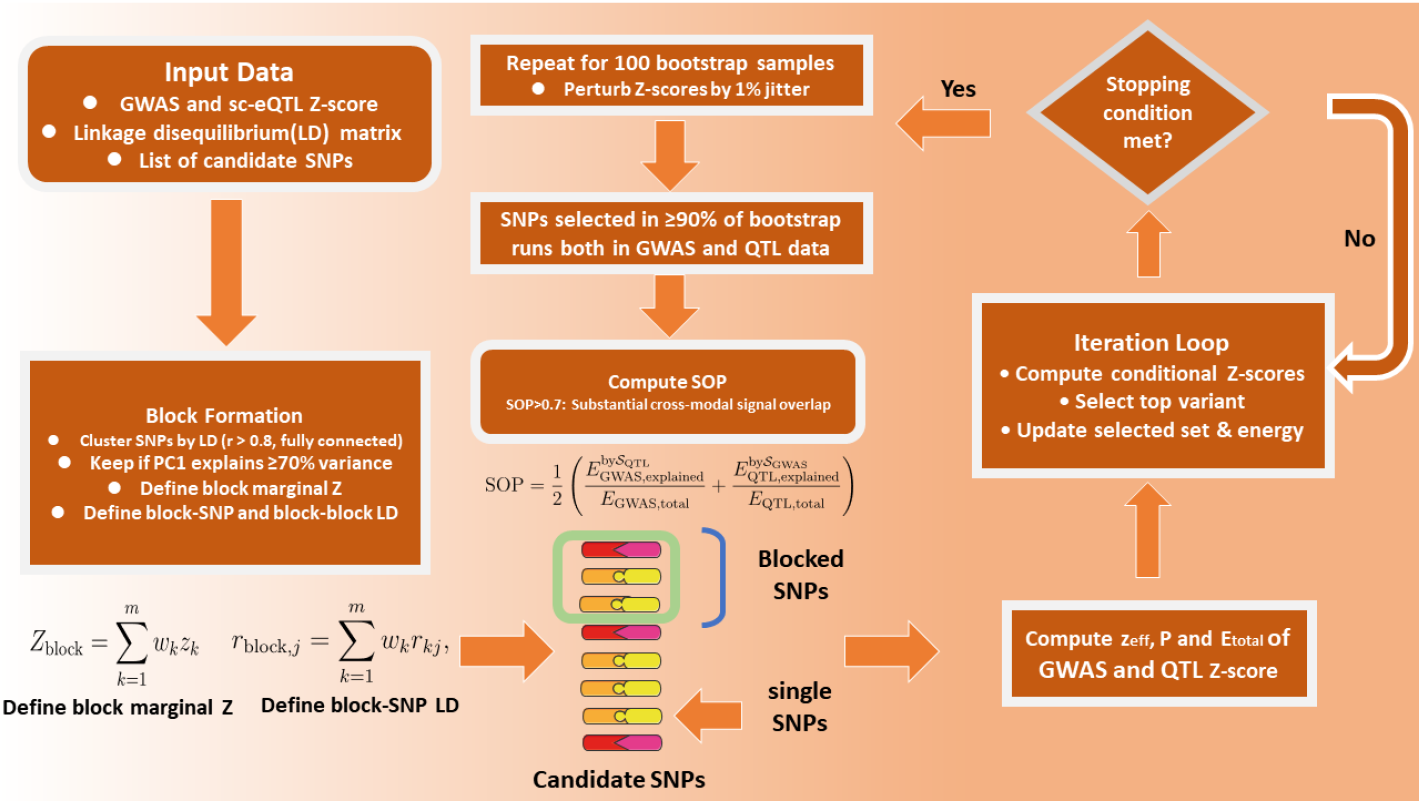
Workflow of the variant prioritization.

“Substantial cross-modal signal overlap” indicates that the genetic associations from GWAS and sc-eQTL analyses show strong reciprocal support within a locus, suggesting a shared underlying causal variant or regulatory mechanism.

### 2. Directional Concordance Between GWAS and sc-eQTL Effects

For each genomic locus satisfying SOP ≥ 0.7, we further assessed the functional directionality of the shared genetic signal by computing a weighted direction agreement score Dt for every gene with significant sc-eQTL support within that locus[24].

Specifically, let Vg denote the set of variants in the locus that are cis-associated with gene g in our sc-eQTL data. For each such gene, we define:

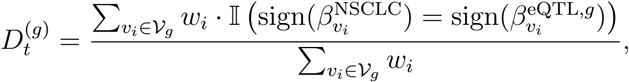

where 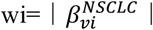 reflects the marginal GWAS effect size of variant vi, and 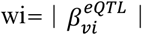 is the effect of vi on the expression of gene. The indicator function I(⋅) equals 1 when the GWAS and eQTL effects are directionally concordant (i.e., both increase or both decrease risk/expression), and 0 otherwise.

This gene-specific score quantifies, within a single high-confidence locus, whether NSCLC risk-increasing alleles tend to upregulate or downregulate each candidate target gene. We interpret Dt(g)≥0.8 as higher NSCLC risk is associated with increased expression of gene g, suggestive of a potential oncogenic role. Conversely, Dt(g)≤0.2 suggests that risk alleles are linked to reduced expression, consistent with tumor-suppressive function. Genes with intermediate scores were considered ambiguous and excluded.

### 3. Locus-Specific Mendelian Randomization comorbidity and NSCLC Risk

To evaluate whether loci prioritized by SOP (SOP ≥ 0.7) harbor genetic architecture consistent with a directional biological link between specific comorbid diseases (idiopathic pulmonary fibrosis (IPF), dermatomyositis, chronic obstructive pulmonary disease (COPD), rheumatoid arthritis (RA), and psoriasis) and NSCLC risk, we performed locus-specific Mendelian randomization using the set of variants identified by our stability-filtered colocalization procedure. For compatibility with Mendelian randomization, we decomposed block-represented signals back to the constituent SNPs. All variants were harmonized to GRCh37 (hg19) and filtered for strand consistency as described in Section 1.1. Non-standard variants (indels, multi-allelic SNPs) were excluded to ensure unambiguous effect direction mapping. We then applied instrument strength filtering (F > 10) to derive a final set of candidate instruments per locus. We then applied Mendelian randomization to serve as a hypothesis-generating evaluation to evaluate consistency of genetic effects across diseases within specific loci[27].

For each SOP locus, we began with the set of bootstrap-stable colocalizated variants and performed formal Mendelian randomization with them. Variants were harmonized to the forward strand, with alleles flipped where necessary to ensure consistent effect direction between exposure and outcome. All variants within each colocalization locus were retained for Mendelian randomization analysis, regardless of individual F-statistics, to preserve the complete posterior probability structure and avoid arbitrary exclusion of potentially causal variants. No LD-based clumping was applied, as SNPs within each locus were intentionally retained to jointly model the regional genetic architecture. For quality control and bias assessment, we calculated and reported both the mean and minimum F-statistic across SNPs within each locus. Loci with mean F-statistic < 10 were flagged for sensitivity analysis to evaluate robustness. We proceeded with causal effect estimation using the inverse-variance weighted method, supplemented by MR-Egger to evaluate robustness. Heterogeneity was assessed using Cochran’s Q statistic, and directional pleiotropy was tested via the MR-Egger intercept. When fewer than three instruments were available, formal MR was not performed.

## Result

### 1. Data resources

Summary-level GWAS data for all outcomes and exposures were selected from studies that only included European ancestral groups as reported in the original publications or metadata. To minimize potential sample overlap between exposure and outcome datasets, we excluded studies sharing overlapping cohorts based on study descriptions and available metadata. Linkage disequilibrium (LD) estimates for European populations were derived from the 1000 Genomes Phase 3 reference panel (EUR) [28].

In total, we incorporated **15** distinct GWAS datasets across six major phenotypes: **2** for chronic obstructive pulmonary disease (COPD), 1 for Idiopathic pulmonary fibrosis (IPF), 1 for Dermatomyositis,**2** for psoriasis, 6 for rheumatoid arthritis, and **4** for Non-small-cell lung cancer. Comprehensive details on GWAS data sources, including sample sizes, phenotype definitions, and data access information, are provided in ***Table 1***. Single-cell eQTL data was drawn from **10** studies, covering **150** different cell types, including monocytes, macrophages (alveolar and interstitial), CD4+ T cells, CD8+ T cells, regulatory T cells, B cells, dendritic cells, NK cells, neutrophils, epithelial cells, and among others. All datasets underwent genomic coordinate conversion and strand harmonization as described in Methods. Single-cell QTL data were obtained from studies in biologically relevant cell types; full details are summarized in ***Supplementary Table 1***.

**Table 1.** Summary of GWAS Data

| Phenotype Definition | Consortium / Study | Sample Size (N) | Ancestry | Data Access |
| --- | --- | --- | --- | --- |
| COPD, Early/Later Onset | IEU | 361194 | European | https://gwas.mrcieu.ac.uk/datasets/ukb-d-COPD_EARLYANDLATER/ |
| COPD | FinnGenR12 | 433208 | European | https://storage.googleapis.com/finngen-public-data-r12/summary_stats/release/finngen_R12_J10_COPD.gz |
| IPF | FinnGenR12 | 52283 | European | https://storage.googleapis.com/finngen-public-data-r12/summary_stats/release/finngen_R12_IPF.gz |
| Dermatomyositis | FinnGenR12 | 494048 | European | https://storage.googleapis.com/finngen-public-data-r12/summary_stats/release/finngen_R12_AB1_DERMATOPHYTOSIS.gz |
| Psoriasis | IEU | 462933 | European | https://gwas.mrcieu.ac.uk/datasets/ukb-b-10537/ |
| Psoriasis | FinnGenR12 | 494941 | European | https://storage.googleapis.com/finngen-public-data-r12/summary_stats/release/finngen_R12_L12_PSORIASIS.gz |
| Rheumatoid Arthritis | IEU | 463010 | European | https://gwas.mrcieu.ac.uk/datasets/ukb-b-11874/ |
| Rheumatoid Arthritis | IEU | 361194 | European | https://gwas.mrcieu.ac.uk/datasets/ukb-d-M13_RHEUMA/ |
| Rheumatoid Arthritis | IEU | 337159 | European | https://gwas.mrcieu.ac.uk/datasets/ukb-a-105/ |
| Rheumatoid Arthritis | IEU | 57284 | European | https://gwas.mrcieu.ac.uk/datasets/ebi-a-GCST002318/ |
| Rheumatoid Arthritis | FinnGenR12 | 331429 | European | https://storage.googleapis.com/finngen-public-data-r12/summary_stats/release/finngen_R12_M13_RHEUMA.gz |
| NSCLC | IEU | 374687 | European | https://gwas.mrcieu.ac.uk/files/ieu-b-4954/ieu-b-4954.vcf.gz |
| NSCLC | IEU | 212453 | European | https://gwas.mrcieu.ac.uk/files/bbj-a-133/bbj-a-133.vcf.gz |
| NSCLC | IEU | 337159 | European | https://gwas.mrcieu.ac.uk/files/ukb-a-54/ukb-a-54.vcf.gz |
| NSCLC | FinnGenR12 | 385195 | European | https://storage.googleapis.com/finngen-public-data-r12/summary_stats/release/finngen_R12_C3_LUNG_NONSMALL_EXALLC.gz |

### 2. Signal-Driven integration of GWAS and sc-eQTL data

We identified 4795 GWAS lead SNPs with p value less than 1 × 10^−6^ that overlapped with at least one significant cis sc-eQTL with nominal p value less than 0.05 across 10 single cell eQTL datasets and 4 NSCLC GWAS datasets. For each lead SNP we defined a ±100 kb candidate locus yielding 4795 genomic regions for analysis as described in Methods. LD information from the 1000 Genomes Phase 3 European panel was available for 100% of variants per locus.

After applying our signal driven prioritization framework we obtained stable variant sets in both GWAS and sc-eQTL data for 803 of the 4795 loci. These 803 loci each yielded a stable variant set in GWAS and a stable variant set in sc-eQTL through independent selection procedures. In these loci the Signal Overlap Proportion SOP which quantifies the degree of shared signal between the two modalities had a mean value of 0.359 (90% confidence interval: 0.342 to 0.376). Notably 258 loci exhibited strong signal concordance with SOP greater than or equal to 0.7. Full details of locus definition, signal prioritization, and directional concordance analyses are provided in the ***Supplementary README and Supplementary Materials(available in 10*.*5281/zenodo*.*17302764)***.

We next assessed directional concordance between GWAS and sc-eQTL effects in the 258 high-SOP loci. Overlapping loci were analyzed independently (Methods1.3), and many corresponded to the same gene or regulatory region. For each gene with significant cis-sc-eQTL support within a locus, we computed a weighted direction agreement score *Dt*(*g*) (Methods, Section 1.3). After filtering for unambiguous directional relationships (*Dt*(*g*)≥0.8 or *Dt*(*g*)≤0.2), we identified 17 distinct cell type–gene pairs (***Table 2***). In each pair, all prioritized variants within the corresponding locus showed consistent effect directions, indicating that NSCLC risk-increasing alleles either uniformly upregulated or downregulated the target gene. Full details of directional concordance are summarized in ***Supplementary Table 2***.

**Table 2.** Distinct cell type–gene pairs with directional concordance (D_t_ ≥ 0.8 or ≤ 0.2) between NSCLC risk and sc-eQTL effects

| Gene | Inferred functional role | Cell type |
| --- | --- | --- |
| ACTN1 | oncogene | CD4 Memory after acquiring effector functions (5 d) |
| CHRNA3 | tumor suppressor gene | Monocyte (Mono) Normal |
| CHRNA5 | tumor suppressor gene | CD4+ Naive T cycling after the first cell division (40h) |
| CHRNA5 | tumor suppressor gene | Monocyte (Mono) Normal |
| CHRNA5 | oncogene | CD4 Memory after acquiring effector functions (5 d) |
| CTSH | oncogene | Monocyte (Mono) |
| HYKK | tumor suppressor gene | CD4 Memory after acquiring effector functions (5 d) |
| IREB2 | tumor suppressor gene | CD4+ Naive T cycling after the first cell division (40h) |
| IREB2 | tumor suppressor gene | Monocyte (Mono) |
| IREB2 | tumor suppressor gene | Monocyte (Mono) Normal |
| MORF4L1 | oncogene | B Cell |
| PSMA4 | oncogene | CD4+ Central Memory T before dividing (16h) |
| PSMA4 | oncogene | CD4+ Naive T cycling after the first cell division (40h) |
| PSMA4 | oncogene | CD4+ Naive T harvested after 16 h stimulation |
| PSMA4 | oncogene | CD4 Memory after acquiring effector functions (5 d) |
| PSMA4 | oncogene | CD4 Memory before dividing (16h) |

Table 2 lists cell type–gene pairs exhibiting strong directional concordance (D_t_ ≥ 0.8 or ≤ 0.2) between NSCLC risk and sc-eQTL effects, along with the inferred functional role of each gene. While some genes show consistent functional assignments across multiple distinct cell types, notably IREB2, which is inferred as a tumor suppressor in CD4+ naive T cells, monocytes, and normal monocytes, and PSMA4, which is consistently classified as an oncogene across diverse CD4+ T cell states, others are supported by evidence from only a single cell type. For example, ACTN1 is inferred as an oncogene in one CD4 memory T cell state, while HYKK and MORF4L1 each yield robust directional inference in a single cell type: HYKK as a tumor suppressor in CD4 memory T cells at 5 days, and MORF4L1 as an oncogene in B cells. These singleton associations still reflect unambiguous functional directionality and align with the reproducible patterns observed for genes like IREB2 and PSMA4.

A notable exception is CHRNA5, which exhibits context-dependent functional inference: it is classified as a tumor suppressor in CD4+ naive T cells cycling after the first division (40 h) and in normal monocytes, but as an oncogene in CD4 memory T cells after acquiring effector functions (5 d). This functional duality underscores that gene roles in NSCLC risk may be highly dependent on cellular context, rather than being universally fixed.

### 3. Locus-Specific Mendelian Randomization

We performed locus-specific Mendelian randomization using genomic loci with SOP score at least 0.7, a threshold indicating robust colocalization between non–small cell lung cancer genome-wide association signals and single-cell expression quantitative trait loci (sc-eQTL) in specific cell types. Only loci with inverse-variance weighted p-value less than 0.05, Cochran’s Q p-value greater than 0.01, and MR-Egger intercept p-value greater than 0.01 were interpreted. Results are shown in ***Table 3***. The full details are provided in the ***Supplementary Table 3***.

**Table 3.**
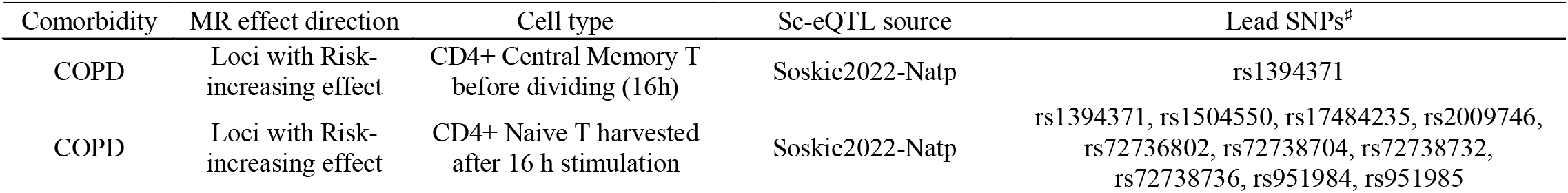
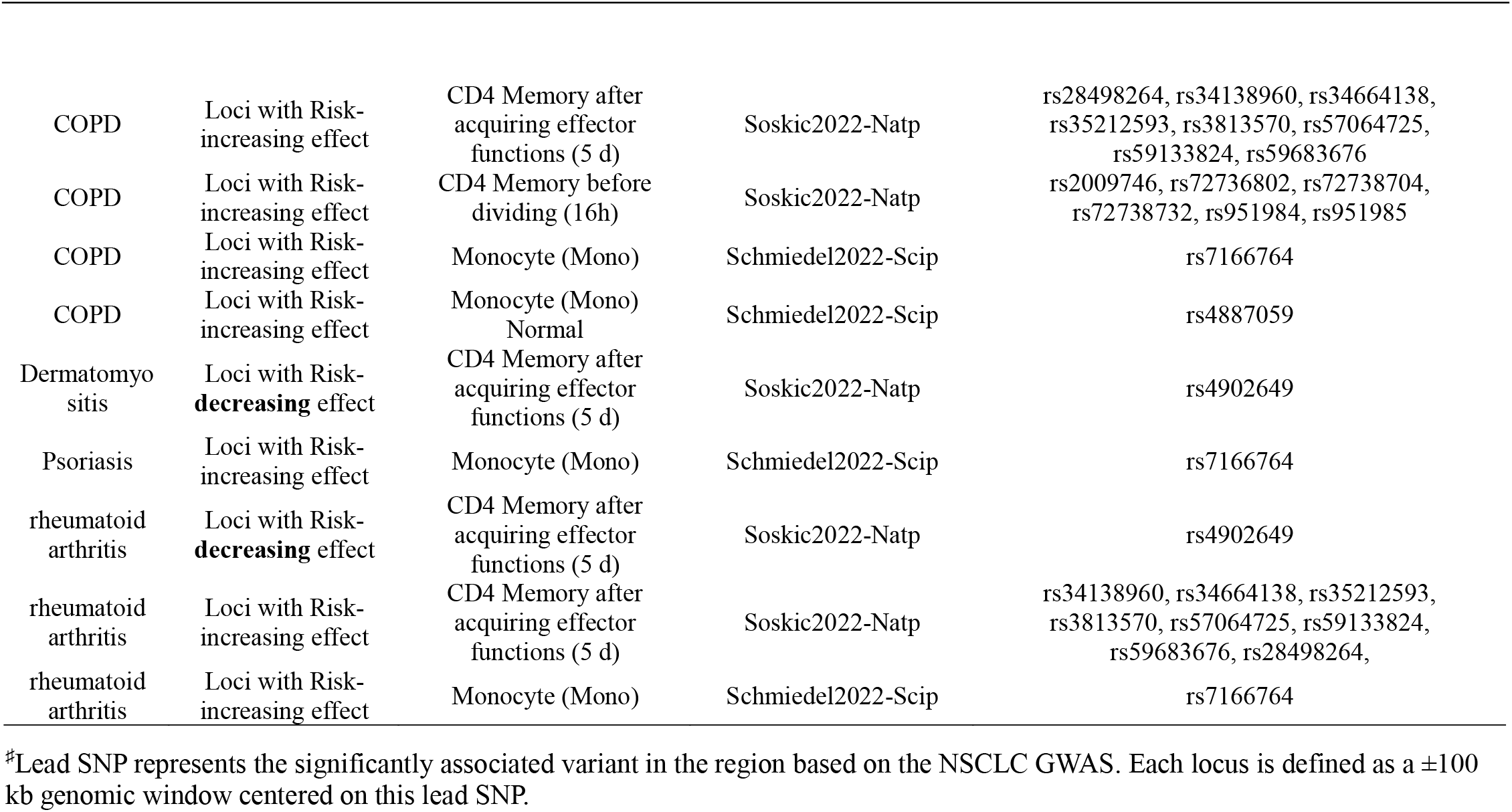
Locus-specific Mendelian randomization supports causal links between comorbidity and NSCLC risk in specific cell type–

| Comorbidity | MR effect direction | Cell type | Sc-eQTL source | Lead SNPs<sup>#</sup> |
| --- | --- | --- | --- | --- |
| COPD | Loci with Risk-increasing effect | CD4+ Central Memory T before dividing (16h) | Soskic2022-Natp | rs1394371 |
| COPD | Loci with Risk-increasing effect | CD4+ Naive T harvested after 16 h stimulation | Soskic2022-Natp | rs1394371, rs1504550, rs17484235, rs2009746, rs72736802, rs72738704, rs72738732, rs72738736, rs951984, rs951985 |
| COPD | Loci with Risk-increasing effect | CD4 Memory after acquiring effector functions (5 d) | Soskic2022-Natp | rs28498264, rs34138960, rs34664138, rs35212593, rs3813570, rs57064725, rs59133824, rs59683676 |
| COPD | Loci with Risk-increasing effect | CD4 Memory before dividing (16h) | Soskic2022-Natp | rs2009746, rs72736802, rs72738704, rs72738732, rs951984, rs951985 |
| COPD | Loci with Risk-increasing effect | Monocyte (Mono) | Schmiedel2022-Scip | rs7166764 |
| COPD | Loci with Risk-increasing effect | Monocyte (Mono) Normal | Schmiedel2022-Scip | rs4887059 |
| Dermatomyositis | Loci with Risk-**decreasing** effect | CD4 Memory after acquiring effector functions (5 d) | Soskic2022-Natp | rs4902649 |
| Psoriasis | Loci with Risk-increasing effect | Monocyte (Mono) | Schmiedel2022-Scip | rs7166764 |
| rheumatoid arthritis | Loci with Risk-**decreasing** effect | CD4 Memory after acquiring effector functions (5 d) | Soskic2022-Natp | rs4902649 |
| rheumatoid arthritis | Loci with Risk-increasing effect | CD4 Memory after acquiring effector functions (5 d) | Soskic2022-Natp | rs34138960, rs34664138, rs35212593, rs3813570, rs57064725, rs59133824, rs59683676, rs28498264, |
| rheumatoid arthritis | Loci with Risk-increasing effect | Monocyte (Mono) | Schmiedel2022-Scip | rs7166764 |

<sup>#</sup>Lead SNP represents the significantly associated variant in the region based on the NSCLC GWAS. Each locus is defined as a $\pm 100$ kb genomic window centered on this lead SNP.

For chronic obstructive pulmonary disease, rheumatoid arthritis, and psoriasis, most loci indicated a risk-increasing effect on non–small cell lung cancer. These loci colocalized with sc-eQTL in immune cell types including CD4+ T cell subsets and monocytes. Across all shared loci(each defined as a ±100 kb genomic window centered on a GWAS lead SNP (p < 1 × 10^−6^)as described in method)the direction of effect on non–small cell lung cancer risk was consistent across comorbid diseases and the immune cell types in which sc-eQTL colocalization was observed. Although some loci associated with different diseases or identified in distinct cell types overlapped, they consistently showed the same direction of effect on non–small cell lung cancer risk. This directional concordance across cell types, diseases, and sc-eQTL datasets reinforces the robustness of these cell-type-specific regulatory relationships in mediating comorbidity-driven susceptibility to lung cancer.

For example, Mendelian randomization in the rheumatoid arthritis, psoriasis, and chronic obstructive pulmonary disease backgrounds, using the locus centered on rs7166764 that colocalized with sc-eQTL in monocytes, consentaneously indicated a causal risk-increasing effect on non–small cell lung cancer. Mendelian randomization in the rheumatoid arthritis and chronic obstructive pulmonary disease backgrounds using eight loci that colocalized with sc-eQTLs in CD4+ memory T cells after 5 days of activation centered on rs34138960, rs34664138, rs35212593, rs3813570, rs57064725, rs59133824, rs59683676, and rs34138960, all similarly indicated consistent causal risk-increasing effects on non–small cell lung cancer. In the same cell type, Mendelian randomization in the dermatomyositis and rheumatoid arthritis backgrounds using the locus centered on rs4902649 that colocalized with an sc-eQTL in CD4+ memory T cells after 5 days of activation indicated a causal protective effect (reduced risk) on non–small cell lung cancer, representing a rare protective signal among the shared loci. Results are shown in Table 3.

## Discussion

Lung cancer remains the leading cause of cancer related mortality worldwide, with a persistently low five-year survival rate largely attributable to late stage diagnosis[2]. While tobacco exposure is the predominant risk factor, a growing body of epidemiological evidence indicates that chronic inflammatory diseases, including chronic obstructive pulmonary disease, psoriasis, and rheumatoid arthritis, are independently associated with increased lung cancer incidence[12–16]. These comorbidities are clinically identifiable and potentially modifiable through immunomodulatory or anti inflammatory interventions. However, the molecular mechanisms underlying these epidemiological associations remain poorly characterized[17–21]. Specifically, it is not yet established which genetic variants, cell types, or biological pathways mediate the observed risk elevation. Without such resolution, efforts to repurpose existing therapies or design preventive strategies for high risk subpopulations remain empirically driven rather than mechanism informed.

Single cell expression quantitative trait locus data offer the potential to connect disease associated genetic variants to gene regulation at cell type resolution. In practice, however, integration of single cell eQTL with GWAS summary statistics is frequently hindered by incomplete or fragmented linkage disequilibrium information. This fragmentation arises from limited sample sizes in single cell studies, sparse coverage of specific cell populations, and technical heterogeneity across sequencing platforms. Conventional colocalization methods, such as HEIDI and COLOC, require a complete sc-QTL data and invertible LD matrix as input[10,29]. When sc-QTL data are missing, the matrix becomes rank deficient and non invertible, causing these methods to fail. Even when LD estimates are available, regions of strong LD, such as a ±100 kb region, often produce ill conditioned matrices with high condition numbers. This numerical instability renders methods that rely on matrix inversion or posterior computation, including SUSIE, unreliable or inapplicable.

We implemented a conditional Z score based iterative selection procedure, conceptually adapted from the COJO framework[22]. The approach operates within local genomic intervals ±100 kb[30]. The method is designed to identify a minimal set of variants that jointly explain the maximum proportion of shared signal between GWAS and single cell eQTL datasets, under conditions of incomplete sc-eQTL data. Candidate variants were first restricted to those overlapping between GWAS and sc-eQTL summary statistics as a prerequisite for cross-modal comparison. However, to ensure robust inference under fragmented LD and heterogeneous study designs, we required substantial shared genetic signal as quantified by the Signal Overlap Proportion (SOP ≥ 0.7).

Our signal-driven integration framework, designed to overcome fragmented variant coverage in single-cell datasets, identified 258 genomic loci exhibiting strong cross-modal concordance (SOP ≥ 0.7) between NSCLC GWAS signals and cell type–specific sc-eQTLs. Within these high-confidence loci, we resolved unambiguous directional relationships between NSCLC risk-increasing alleles and target gene expression across 17 distinct cell type–gene pairs, enabling functional annotation of non-coding risk variants with single-cell resolution.

ACTN1 was inferred as an oncogene in CD4 memory T cells, a finding that aligns with laboratory evidence from head and neck squamous cell carcinoma where ACTN1 promotes tumor growth and chemoresistance through β-catenin signaling[31]. CTSH was uniformly classified as an oncogene in monocytes, compatible with epidemiological data showing that genetically predicted higher CTSH expression is associated with increased lung cancer risk, likely reflecting its role in alveolar epithelial biology[32]. PSMA4 was robustly annotated as oncogenic in multiple CD4^+^ T cell subsets, in agreement with functional studies in lung cancer cell lines demonstrating that PSMA4 supports proliferation and suppresses apoptosis via proteasome regulation[33]. IREB2 was assigned tumor-suppressive roles in both CD4^+^ T cell subsets and monocytes, a directionality supported by clinical observations in kidney cancer where high IREB2 expression correlates with better survival, enhanced ferroptosis, and greater immune infiltration[34]. MORF4L1 was identified as an oncogene in B cells, consistent with its overexpression in hepatocellular carcinoma and its identification as a CRBN substrate involved in chromatin remodeling, though again in malignant epithelial cells[35]. CHRNA3 was assigned a tumor-suppressive role in monocytes in our study, and large-scale GWAS and clinical association studies have consistently linked SNPs in or near CHRNA3 with increased lung cancer susceptibility and altered response to chemotherapy, underscoring the biological relevance of this genomic region, although no direct functional evidence currently defines the directionality or cellular mechanism of CHRNA3 itself[36].

A particularly exceptional observation was the context-dependent functional role of CHRNA5. In effector-memory CD4^+^ T cells, NSCLC risk alleles were associated with reduced CHRNA5 expression, supporting an oncogenic interpretation, whereas risk alleles correlated with increased CHRNA5 expression in naive cycling CD4^+^ T cells and monocytes, consistent with a tumor-suppressive function. These opposing directional effects occur despite the fact that existing literature on CHRNA5 is limited to statistical associations between its genetic variants and lung cancer susceptibility or smoking behavior, with no experimental validation of its causal role or directionality in specific cell types[33,37]. Our findings therefore propose a mechanistic model hypothesis, wherein CHRNA5 exerts divergent functions depending on immune cell state. This highlights that cellular context can fundamentally reshape the interpretation of cancer risk alleles, a principle essential for advancing precision oncology beyond bulk-tissue paradigms.

Beyond gene regulation, locus-specific Mendelian randomization lent causal support to epidemiological links between inflammatory comorbidities and NSCLC. Our locus-specific Mendelian randomization analysis supports a causal role of inflammatory comorbidities in non-small cell lung cancer susceptibility in specific locus. Genetic liability to chronic obstructive pulmonary disease, rheumatoid arthritis and psoriasis was consistently associated with increased NSCLC risk at shared genomic loci that colocalize with single-cell eQTLs in immune cell types such as monocytes and activated CD4+ memory T cells. Notably, the locus centered on rs7166764, which shows regulatory effects in monocytes, demonstrated a concordant risk-increasing effect on NSCLC across all three diseases. Similarly, eight different loci centered on rs34138960, rs34664138, rs35212593, rs3813570, rs57064725, rs59133824 or rs59683676, all colocalizing with eQTL signals in CD4+ memory T cells after 5 days of activation, showed consistent risk-increasing effects in both rheumatoid arthritis and chronic obstructive pulmonary disease backgrounds.

Interestingly, the locus concentrated on rs4902649, colocalizing with eQTL signals in activated CD4+ memory T cells, stood out as an exception, where genetic predisposition to dermatomyositis and rheumatoid arthritis was associated with reduced NSCLC risk. This rare protective signal suggests that not all immune-related genetic variants promote cancer development. The observation that multiple distinct inflammatory diseases converge on the same SNPs within defined immune cell types and show coordinated effects on lung cancer risk points to highly specific regulatory mechanisms that transcend individual diagnoses. These findings highlight the value of integrating cell type-resolved functional genomics with genetic epidemiology to uncover shared pathways in inflammation-associated carcinogenesis.

Our study has several important limitations. First, the validity of our signal-driven inference relies on the assumption that unobserved common variants are adequately captured through linkage disequilibrium with observed, nominally significant SNPs in dense, high-LD regions. While this may hold when the observed set is sufficiently large and well distributed, it is not guaranteed, particularly because accurate proxy representation requires not only correlation in genotype (LD) but also concordance in effect size direction and magnitude across traits. In sparse single-cell eQTL data, this dual requirement is difficult to verify, and missing variants with distinct regulatory effects could distort signal reconstruction. Thus, our approach provides a conditional approximation under idealized assumptions rather than a robust solution to data fragmentation. Second, the sc-eQTL datasets used in this study were derived from non-cancerous or non-lung contexts, which may not reflect the gene regulatory landscape relevant to NSCLC pathogenesis. Finally, our framework identifies statistical concordance between GWAS and sc-eQTL signals but does not establish causality. Even locus-specific Mendelian randomization serves here only as a consistency check across traits, not as proof of a causal chain. All findings should therefore be interpreted as correlational hypotheses requiring experimental validation[10].

## Conclusion

Integrative analysis of GWAS and single-cell eQTL data enables the identification of shared regulatory architectures that link inflammatory comorbidities to lung cancer risk. This framework highlights the potential role of immune cell-type-specific gene regulation in disease etiology and may inform future strategies for risk stratification and mechanistic investigation.

## Data Availability

All data produced in the present study are available upon reasonable request to the authors
manuscript has been seen and approved by all authors.

