## Supplementary Methods for "Integrative Analysis of GWAS and Single-Cell eQTL Data Identifies Immune Cell-Type-Specific Genetic Signals Co-occurring in Inflammatory Diseases and Lung Cancer Risk"

### Supplementary Methods: Signal-Driven integrative variant prioritization under Fragmented LD

Yu Langxuan

October 8, 2025

#### A. Block Formation

To improve representation in high-linkage disequilibrium (LD) regions and reduce redundancy within a candidate locus, we grouped SNPs into non-overlapping functional blocks based on pairwise LD Specifically, SNPs with pairwise correlation ($r > 0.8$) were clustered into blocks such that all members within a block are mutually highly correlated. In cases where a SNP could belong to multiple overlapping blocks, it was assigned to the block with the higher mean within-block $r$. Within each block, the first principal component (PC1) of the member SNPs' LD matrix was computed and used as a "pseudo-SNP" to represent the shared genetic signal. A block was retained only if PC1 explained $\geq 70\%$ of the variance among its members; otherwise, SNPs with low PC1 loadings were iteratively removed until the criterion was met or fewer than two SNPs remained. This approach ensures that each block captures a coherent regulatory unit for downstream analysis.

Each valid block was represented by its first principal component (PC1), which captures the dominant shared signal among member SNPs. The representative Z-score of the block, denoted $Z_{\text{block}}$, was computed as a weighted average of the marginal Z-scores of its member SNPs:

$$Z_{\text{block}} = \sum_{k=1}^{m} w_k z_k$$

where $w_k$ is the loading of SNP $k$ on PC1, and $z_k$ is its marginal Z-score.

To ensure that $Z_{\text{block}}$ has unit variance under the null, the loadings were explicitly normalized:

$$w_k \leftarrow \frac{w_k}{\sqrt{\sum_{i,j} w_i w_j r_{ij}}}$$

such that $\text{Var}(Z_{\text{block}}) \approx 1$, making it approximately standard normal under H0.

The LD between a block and an external SNP $j$ was defined as the expected correlation between the block's PC1 and SNP $j$:

$$r_{\text{block},j} = \sum_{k=1}^{m} w_k r_{kj},$$

where $r_{kj}$ is the pairwise LD (correlation) between SNP $k$ in the block and external SNP $j$. This represents the linear projection of SNP $j$ onto the dominant shared signal of the block.

For two blocks A and B, their inter-block LD was computed as a regularized Pearson-like correlation:

$$r_{A,B} = \frac{\sum_{i\in A}\sum_{j\in B} w_i^{(A)} w_j^{(B)} r_{ij}}{\sigma_A \sigma_B}.$$

Where (i) $w_i^{(A)}, w_j^{(B)}$: PC1 loadings in blocks A and B; (ii) $r_{ij}$: LD between SNP $i$ and $j$; (iii) $\sigma_A = \sqrt{\sum_{i,j\in A} w_i^{(A)} w_j^{(A)} r_{ij}}$: empirical standard deviation of PC1 in block A. This generalizes the block-to-SNP LD to block-level interactions.

### B. Conditional Z-Score-Based Fine-Mapping

To identify compact and stable sets of genetic variants that collectively explain the shared signal between GWAS and sc-eQTL associations in the presence of fragmented sc-eQTL summary statistics, we implemented a deterministic, iterative selection procedure based on conditional Z-scores. This approach avoids Bayesian matrix inversion, which can be unstable when the available sc-eQTL signals are sparse and non-representative of the full cis-regulatory landscape. Instead, it sequentially selects the variant or variant block that accounts for the largest residual shared signal, constructing a minimal set that captures the majority of the association evidence. Stability is assessed via bootstrap resampling to ensure robustness to perturbations in effect size estimates and incomplete variant coverage.

Firstly, to quantify the total association signal in a locus, we define a signal energy metric based on the quadratic form. The total shared signal between GWAS and sc-eQTL associations is quantified as a regularized quadratic form:

$$E_{\text{total}} = \mathbf{z}_{\text{eff}}^T \mathbf{P} \, \mathbf{z}_{\text{eff}}$$

where $\mathbf{z}_{\text{eff}}$ is the background-variance-corrected marginal Z-score vector, and $\mathbf{P} = \mathbf{U}\boldsymbol{\Lambda}^{-1}\mathbf{U}^T$ is the projection operator derived from spectral truncation of the LD matrix $\mathbf{R} = \mathbf{U}\boldsymbol{\Lambda}\mathbf{U}^T$. Only eigenvalues $\lambda_i \geq \max(0.2, 10^{-6}\lambda_{\max})$ are retained to ensure numerical stability and robustness to noise.

Then, at each iteration $k$, the variant or SNP block with the largest absolute conditional Z-score was selected:

$$j^* = \arg\max_j \left| Z_j^{(k-1)} \right|$$

where $\mathbf{Z}_j^{(k-1)}$ is the partially conditioned Z-score of variant $j$ at step $k-1$, computed by regressing out the effects of all previously selected variants using the LD structure. Specifically, after selecting $j^*$, the updated conditional Z-score vector is recomputed as:

$$\mathbf{z}_{\text{cond}}^{(k)} = \mathbf{z} - \mathbf{R}_{\cdot,S_k} \left(\mathbf{R}_{S_k,S_k}\right)^+ \mathbf{z}_{S_k}$$

where $\mathbf{R}_{S_k,S_k}^+$ denotes the Moore-Penrose pseudoinverse of the LD submatrix among selected variants. This update follows the COJO framework, where the conditional Z-scores are obtained by regressing out the marginal effects of selected variants using the LD structure, which ensures accurate modeling of local LD patterns in the original genetic space.

To maintain numerical stability and enable direct comparison across variants, all energy contributions are computed in a spectrally truncated principal component space derived from the global LD matrix. For each variant $j$, its contribution to the shared signal is quantified as:

$$E_j = \mathbf{z}_j^T \mathbf{P} \, \mathbf{z}_j$$

where $\mathbf{z}_j$ is the Z-score vector of the variant or block (e.g., PC1 score), projected into the same spectrally truncated space using consistent background correction and $\mathbf{P}$ operator. This ensures that all contributions are measured on a common scale, enabling direct comparison and summation.

The cumulative explained energy is computed as the difference between the total signal energy and the residual energy in the spectrally truncated space:

$$E_{\text{explained}}^{(k)} = \mathbf{z}_{\text{eff}}^T \mathbf{P} \mathbf{z}_{\text{eff}} - \mathbf{z}_{\text{cond}}^{(k)T} \mathbf{P} \mathbf{z}_{\text{cond}}^{(k)}$$

which ensures that the explained energy reflects the true joint contribution of selected variants, accounting for linkage disequilibrium. The signal completion metric is then defined as $C_k = E_{\text{explained}}^{(k)} / E_{\text{total}}$, measuring the proportion of total signal explained by the current set. The procedure terminates when:

$$\text{If } \frac{E_{\text{explained}}^{(k)}}{E_{\text{total}}} \geq 0.8 \quad \text{or} \quad \max \left| \mathbf{Z}_{\text{cond}}^{(k)} \right| < 1.645, \quad \text{then stop.}$$

All energy estimates are computed within the same spectrally truncated subspace, ensuring that signal contributions are evaluated on a common scale.

To ensure the reliability of selected variants and blocks, we perform bootstrap resampling with 100 iterations, each time applying a 1 percent random perturbation to the Z scores. Within each replicate, we identify the minimal set of variants or blocks that explains at least 90 percent of the projected signal. A variant or block is included in the final output only if it appears in at least 90 percent of the bootstrap replicates. This results in a compact and stable set of SNPs or blocks that consistently represent the shared GWAS and sc-eQTL signal across perturbations.
