## Supplementary Notes for "Integrative Analysis of GWAS and Single-Cell eQTL Data Identifies Immune Cell-Type-Specific Genetic Signals Co-occurring in Inflammatory Diseases and Lung Cancer Risk"

### Supplementary Note: A Framework for Cross-Trait Signal Integration under Fragmented LD

Yu Langxuan

This note outlines the statistical framework underlying our refined spectral fine-mapping pipeline. We integrate established techniques to improve stability and interpretability in regions of high linkage disequilibrium (LD), allelic heterogeneity, and incomplete variant coverage.

The focus is practical robustness. We apply spectral thresholding at $\tau = \max(0.2, 10^{-6}\lambda_{\max})$ to suppress noise amplification in ill-conditioned LD matrices of a ±100 kb region. Within this stabilized subspace, we define a signal completion metric $C_k$ that quantifies the proportion of total signal explained by selected variants. $C_k$ increases monotonically with inclusion of new SNPs or blocks and naturally saturates as coverage improves, providing a data-driven criterion for assessing signal recovery.

These methods build upon existing approaches, but we emphasize their integration into a coherent workflow for refining signals around candidate genes, particularly in our previously described pipeline where variants may be incomplete. The resulting stable SNP sets are intended to serve as inputs for downstream analyses, ensuring that observed signal shifts across molecular traits reflect biological differences.

We present basic theoretical properties (e.g., bounded increment, consistency under convergence) not as proofs of superiority, but as sanity checks that the procedure behaves as intended under realistic modeling assumptions. This is especially important in the context of GWAS-QTL colocalization, where consistent and stable variant selection is critical for reliable causal inference.

#### A. Unified Definition of SNP- and Block-Level LD

Let $g_i$ denote the standardized genotype of SNP $i$. For a genomic block $A$, its first principal component (PC1) is defined as:

$$\mathrm{PC}_A = \sum_{i \in A} w_i^{(A)} g_i, \quad \text{with} \quad \sigma_A^2 = \mathrm{Var}(\mathrm{PC}_A) = \sum_{i,j \in A} w_i^{(A)} w_j^{(A)} r_{ij}.$$

The block-to-block LD between $A$ and $B$ is then:

$$r_{A,B} = \frac{\sum_{i \in A, j \in B} w_i^{(A)} w_j^{(B)} r_{ij}}{\sigma_A \sigma_B} = \mathrm{Corr}(\mathrm{PC}_A, \mathrm{PC}_B),$$

which is the Pearson correlation between the two PC scores. This generalizes naturally to block-to-SNP LD:

$$r_{A,j} = \sum_{i \in A} w_i^{(A)} r_{ij},$$

the weighted average LD from block $A$ to SNP $j$. Thus, the same principle applies consistently across SNP-SNP, SNP-block, and block-block levels, ensuring dimensional and interpretational coherence.

#### B. Signal Completion Metric and Effective Monotonicity

Let $\mathbf{R}$ be the LD matrix over a candidate region, and let $\hat{\mathbf{z}}$ be the vector of background-corrected marginal Z-scores from association testing. In our small region (±1Mb), the matrix $\mathbf{R}$ is often ill-conditioned, making direct inversion unstable. To address this, we regularize the geometry by projecting signals onto a well-behaved subspace:

$$\mathcal{V}_\tau = \operatorname{span}\{\mathbf{v}_i : \lambda_i \geq \tau\}, \quad \tau = \max(0.2, 10^{-6}\lambda_{\max}(\mathbf{R})).$$

This retains only the principal components of $\mathbf{R}$ with sufficiently large eigenvalues, filtering out noise-dominated directions. Let $\mathbf{P}_{\geq\tau}$ denote the orthogonal projector onto $\mathcal{V}_\tau$, and let $\mathbf{R}_\tau^\dagger$ be the pseudoinverse of $\mathbf{R}$ restricted to this subspace.

Given a selected set of variants $\mathcal{S}_k$, we compute the residual Z-score vector $\hat{\mathbf{z}}_k^{(\mathrm{res})}$ after conditioning on $\mathcal{S}_k$, using the regularized LD structure. The **signal completion metric** is then defined as:

$$C_k = 1 - \frac{(\hat{\mathbf{z}}_k^{(\mathrm{res})})^T \mathbf{P}_{\geq\tau} \hat{\mathbf{z}}_k^{(\mathrm{res})}}{\hat{\mathbf{z}}^T \mathbf{P}_{\geq\tau} \hat{\mathbf{z}}}, \quad \text{provided } \hat{\mathbf{z}}^T \mathbf{P}_{\geq\tau} \hat{\mathbf{z}} > 0.$$

This measures the proportion of total signal energy explained by $\mathcal{S}_k$, based on the reduction in residual signal under the regularized geometry. It behaves similarly to an $R^2$ statistic in regression, quantifying how well $\mathcal{S}_k$ "accounts for" the observed associations.

When a new SNP $s_{\mathrm{new}}$ is added, its contribution is assessed via the residual Z-score:

$$z_{\mathrm{res}} = z_{\mathrm{new}} - \mathbf{r}_{\mathrm{new},\mathcal{S}_k}^T \mathbf{R}_{\mathcal{S}_k}^\dagger \mathbf{z}_{\mathcal{S}_k},$$

which captures the part of its signal not predictable from the current set. The increase in explained energy is proportional to $z_{\mathrm{res}}^2$ times a positive weight $q_{\mathrm{new}}$, derived from the local curvature of the regularized model (related to the Schur complement). Since both factors are non-negative, this ensures that $C_k$ is effectively monotonic, which increases with meaningful additions and plateaus when no further independent signal is found.

Small numerical fluctuations may cause minor dips in $C_k$, but are suppressed by the spectral thresholding ($\tau$), which stabilizes computations and avoids overfitting to poorly estimated directions.

### C. Signal Saturation, Estimation Stability, and Cross-Trait Consistency Evaluation

Let $\mathcal{S}_k$ denote the set of variants selected from the available (and typically incomplete) sc-eQTL summary statistics within a cis-region, and let $C_k$ be the signal completion metric, which is non-decreasing as additional variants are incorporated into $\mathcal{S}_k$. We interpret $C_k \geq 0.90$ as indicating substantial recovery of the shared GWAS–sc-eQTL signal, and $C_k \geq 0.99$ as suggesting local saturation.

The stability of the estimator $\hat{C}_k$ depends on sample size $n$. As $n \to \infty$, we have $\hat{\mathbf{z}}_n \xrightarrow{p} \mathbf{z}$ and $\hat{\mathbf{R}}_n \xrightarrow{p} \mathbf{R}$, implying that the estimated signal subspace $\hat{\mathcal{V}}_{\tau_n}$ converges in probability to the true signal subspace $\mathcal{V}_{\mathrm{signal}}$. By the continuous mapping theorem, it follows that $\hat{C}_k \xrightarrow{p} C^*$, where $C^*$ is the population-level signal completion. Crucially, the same projection operator $\mathbf{P}_{\geq\tau}$ and spectral threshold $\tau$ must be applied consistently in both the numerator and denominator of $C_k$; otherwise, comparisons across estimates become invalid.

This assumption is supported by the use of high-quality reference panels (e.g., 1000 Genomes Phase 3 EUR), which provide accurate LD estimates for common variants, and by the block-based aggregation strategy, which enhances robustness to individual SNP omission through principal component summarization.

This property enables preliminary signal evaluation using a smaller SNP set. Conclusions remain approximately valid after expanding the number of variants. Grouping variants into blocks further improves stability by reducing sensitivity to individual SNP inclusion.

In multi-omics analyses, the same variant set $S$ can be evaluated across molecular phenotypes using $C(S; \mathbf{z}^{(1)})$ and $C(S; \mathbf{z}^{(2)})$. If both values are high and the projected signals $\mathbf{P}_{\geq\tau}\mathbf{z}^{(1)}$ and $\mathbf{P}_{\geq\tau}\mathbf{z}^{(2)}$ are highly correlated, this suggests potential shared regulatory architecture. This observation does not imply statistical colocalization, but may serve as a pre-screening indicator.

*Note:* The derivations above aim to clarify the behavior and assumptions of the proposed pipeline. They are not claimed as theoretical novelty, but as justification for practical use. We welcome feedback on potential improvements.
